# Non-invasive epidermal proteomics and machine learning permits molecular subclassification of psoriasis and eczematous dermatitis

**DOI:** 10.1101/2024.09.24.24314282

**Authors:** Yiwei Wang, Michael J. Murphy, Tingting Chen, Gloria Chen, Princess Edemobi, Muhammad H. Junejo, A. Mitchel Wride, Sarah L. Spaulding, Kenneth Y. Tsai, Jeffrey M. Cohen, William Damsky

**Author notes:** **Corresponding authors:** William Damsky, MD, PhD, Department of Dermatology and Pathology, Yale School of Medicine, 333 Cedar St, LCI 501, PO Box 208059, New Haven, CT 06510., Jeffrey M. Cohen, MD, MPH, Department of Dermatology, Yale School of Medicine, 333 Cedar St, LCI 501, PO Box 208059 New Haven, CT 06510. Yiwei Wang and Michael J. Murphy contributed equally to this work.

## Abstract

Current approaches to selecting molecularly targeted therapies (biologics and oral small molecules) for immune-mediated skin diseases largely overlook interindividual immunologic heterogeneity, in part due to challenges of sample collection and the lack of broadly accepted biomarkers of therapeutic response. We sought to develop a rapid, minimally invasive method for obtaining and measuring biomarkers from skin and to generate predictive models of therapy response in two common inflammatory skin diseases, psoriasis and eczema. Here we present Detergent-based Immune Profiling System (DIPS), which enables painless and non-scarring collection of full thickness epidermal protein from skin and is suitable for downstream 45-plex immune protein biomarker analysis. We first developed machine learning models with the goal of accurately distinguishing between eczema (n=55) and psoriasis (n=74). Subsequently, models that correlate biomarker patterns with treatment response or nonresponse to commonly used biologic therapies were developed. We subsequently developed DIPS-Derm, a web-based platform that provides automated diagnostic and treatment predictions from data generated with DIPS. These results support the promise of artificial intelligence (AI)-driven precision dermatology and highlight the clinical potential of DIPS for personalized medicine in inflammatory skin disease.

## Introduction

Psoriasis and eczema are among the most common inflammatory skin conditions ^1^. These diseases are increasingly treated with molecularly targeted therapies. Approved molecular targets in psoriasis include TNFα, interleukin (IL)-12/23 (p40), IL-23 (p19), IL-17A, IL17-A/F, IL-17R, IL-36R, tyrosine kinase 2 (TYK2), phosphodiesterase 4 (PDE4), and aryl hydrocarbon receptor (AhR). In eczema, approved targets include IL-4Rα, IL-13, IL-31, Janus kinase (JAK)1, PDE4, and AhR, with others such as OX40/OX40L agents showing promise in clinical trials. Outside of specific clinical scenarios, and generally based on medical comorbidities, the mechanism of action chosen for any individual patient is generally empiric, based on population-level efficacy, and importantly without incorporation of individualized molecular data. Over time, the task of selecting the appropriate medication and the potential for differential outcomes based on mechanism of action will only increase as more molecularly-targeted therapies are developed for these and other inflammatory skin diseases.

In clinical practice, it is not uncommon for patients with either psoriasis or eczema to have a suboptimal response to one targeted therapy but then an excellent response to another ^2, 3^. Roughly 10-35% of patients with psoriasis and 35-60% of patients with eczema do not achieve clear or almost clear skin with common first-line therapies such as IL-23 inhibition for psoriasis and dupilumab (IL-4Rα) for eczema ^4, 5^. Occasionally, there may even be exacerbation of the underlying disease, or morphology switching, if the wrong medication is selected ^6, 7^. In a real-world analysis of eczema patients treated with four common systemic therapies, 51% of patients switched from the first line biologic to another therapy within 1 year ^8^.

It has become increasingly apparent that within each clinical diagnosis, eczema and psoriasis, there is intra-disease immunologic heterogeneity ^9^. Further, occasional cases of psoriasis may show eczematous features and vice-versa, leading to misdiagnoses or inconclusive diagnoses, even if skin biopsies, the current gold-standard in diagnosis, are performed ^9^. This variability has led some investigators to divide each disease conceptually into distinct disease endotypes based on differing clinical manifestations, molecular profiles, and/or host characteristics such as gender, race, and age ^9, 10, 11^. Skin biopsy cannot differentiate among disease endotypes.

Such disease endotypes and their underlying immunologic variability, have been proposed as key drivers of variable responses to the same targeted therapy between individual patients with the same diagnosis. In eczema, while most patients have at least some degree of a Type 2 (Th2) immune activation (IL-4, IL-13, IL-31), other patients may have concomitant or even predominant Type 1 (Th1), Type 3 (Th17), and/or Th22 activation ^12, 13, 14, 15^. Psoriasis subtypes may also exhibit distinct immunology ^9, 13^; for example, in pustular psoriasis, an innate immune driver (IL-36) appears to be predominant, as opposed to an adaptive Th17 polarized response in classic psoriasis vulgaris ^16^.

Assessing the predominant immunologic drivers has been shown to correlate with treatment response or non-response in some instances in patients with these diseases ^13, 14, 15, 17^, but these approaches are generally not feasible in most clinical settings. Thus, treatment selection in both diseases remains largely empiric, without the ability to dissect the unique immunology of any individual patient to inform therapy selection in a rapid, clinically relevant fashion.

The ideal test for clinically relevant immunologic profiling in inflammatory skin diseases would be easy and rapid to obtain, painless and non-scarring, accurate, and have the potential for short, clinically relevant turnaround. We developed and evaluated a new approach we term Detergent-based Immune Profiling System (DIPS). DIPS enables minimally invasive collection of epidermal proteins from full thickness epidermis for downstream 45-plex immune protein biomarker measurement. We performed DIPS on patients with psoriasis and eczema, as well as normal skin from healthy control patients. We developed diagnostic and treatment-response biomarker models for eczema and psoriasis and built machine learning classifiers with high predictive performance. Last, we created a web-based platform, DIPS-Derm (https://dips-derm.onrender.com), that is accessible for research use and serves as a foundation for further development of this approach for AI-driven precision dermatology.

## Results

### Study design

The study was conducted in six stages: 1) development and optimization of the DIPS approach, 2) sample collection in patients with psoriasis and eczema, 3) multiplexed protein biomarker analysis on the samples, 4) model development with classifiers to distinguish eczema from psoriasis, 5) model development for treatment response pattern classifiers within each disease, and 6) development and deployment of a web-based platform (**Fig. 1A**). Model training and internal validation were performed in the discovery cohort, with validation conducted across several independent datasets. The final models were implemented within our web-based clinical decision support platform, DIPS-Derm (<u>D</u>etergent-based Immune <u>P</u>rofiling <u>S</u>ystem for <u>Derm</u>atology), enabling diagnostic classification and/or treatment response predictions based on user-submitted DIPS-based protein biomarker data (**Fig. 1A**).

**Fig. 1.**
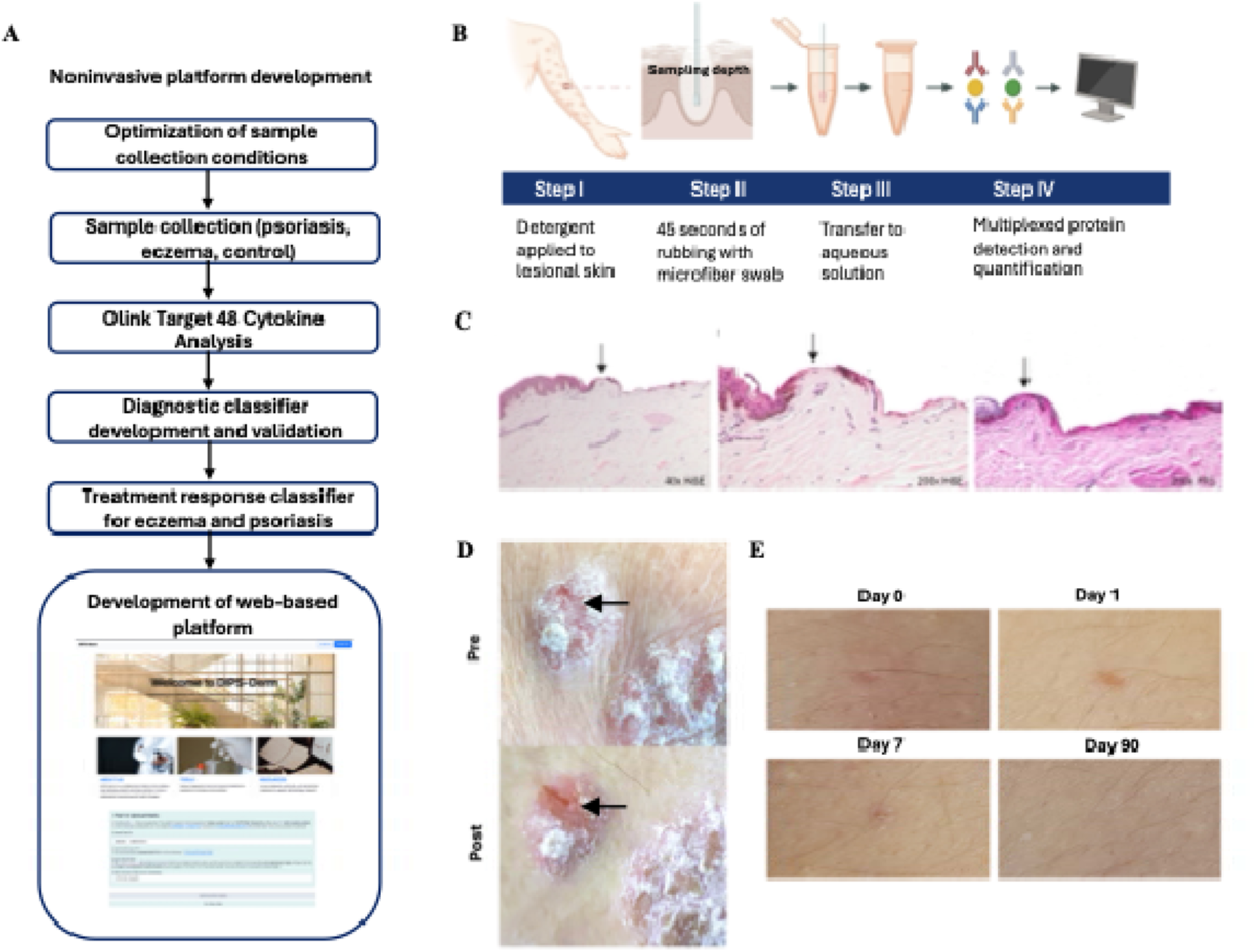
Development and validation of DIPS. **(A)** Overall study design. **(B)** Schematic of the DIPS protocol. **(C)** Histologic evaluation of *ex vivo* skin samples post-collection confirms full thickness sampling of the epidermis. H&E (40× and 200×) and PAS (20×) staining. **(D)** Representative lesional skin images before (pre) and after (post) DIPS sample collection in a patient with psoriasis. The arrow indicates the area of sampling. **(E)** Representative images of the DIPS collection site in a healthy control patient (days post collection).

### Development and evaluation of DIPS

We first focused on development of a noninvasive method that would allow for rapid collection of protein directly from skin to be used for subsequent downstream biomarker analysis at the protein level. We selected Clearista Retexturizing gel, which was initially developed for protein collection from skin ^18, 19^ but has since been shown to also enable DNA isolation ^20^. It has more recently been pursued commercially only for aesthetic purposes. Clearista is based on a combination of two nondenaturing surfactants, 0.25% Laureth-4 and 0.25% DPS-30. We first evaluated the ability of Clearista to solubilize human epidermis *ex vivo* when applied to and rubbed against intact human skin with a small applicator (**Fig. 1B**). Among six tested applicators, a microfiber-tipped cosmetic swab was selected based on its superior efficiency in solubilizing full-thickness epidermis over a small area (∼2 × 1 mm) and minimizing sample loss during transfer of the sample to phosphate buffered saline (PBS) (data not shown). We added protease inhibitors to the PBS to improve protein stability. Histological examination of the *ex vivo* skin after sample collection was used to determine that a 45-second swabbing duration was optimal for achieving full-thickness epidermal solubilization while preserving the dermal-epidermal junction, as confirmed by Periodic acid–Schiff (PAS) staining (**Fig. 1C**).

Next, we evaluated this approach in healthy controls and patients with psoriasis. As with the *ex vivo* skin, sample collection resulted in a visible superficial erosion and transient depression of the skin in the sampled area (**Fig. 1D**). DIPS was very well-tolerated as assessed using an established Local Tolerability Score metric ^21^ **(Table S1**). We also found that re-epithelialization was rapid and occurred without scarring or pigmentary alteration (**Fig. 1E**).

To assess the feasibility and accuracy of downstream protein biomarker measurement using samples generated with this approach, we analyzed the samples using Olink Target 48 Cytokine panel, which provides absolute quantification (pg/μL) of 48 relevant cytokines/chemokines. Target 48 Cytokine includes cytokines and chemokines relevant to overall immune polarization patterns including Type 1 (IFN-γ, CXCXL9, CXCXL10, and TNFα), Type 2 (IL-4 and IL-13), and Type 3 (IL-17A and IL-17F), as well as others. Ultimately, protein was successfully quantified from samples collected from 74 individuals with psoriasis (PS), 55 with eczema (AD), and 22 healthy controls (HC) without inflammatory skin disease.

We were reassured to see that IL-17A/IL-17F were among the most upregulated markers in psoriasis, while IL-4 and IL-13 were among the most upregulated markers in eczema (**Fig. S1 and Fig. S2**). Negligible expression of these markers was observed in the healthy controls. A three-dimensional NMDS plot was generated to visualize the differential protein abundance patterns across the three groups and revealed distinct protein expression signatures that led to significantly different clustering of AD, PS, and HC samples (p = 0.001, **Fig. S1C**).

We termed this overall approach, <u>D</u>etergent-based Immune <u>P</u>rofiling <u>S</u>ystem (DIPS). We found that DIPS enables efficient epidermal protein capture while preserving disease-relevant molecular information, supporting its further development for epidermal proteomics and biomarker quantification in inflammatory skin diseases.

### Diagnostic Classifier Development for Psoriasis and Eczema

We next aimed to develop a model that could accurately distinguish eczema and psoriasis using the biomarker data from DIPS. Model development was performed using a machine learning-based feature importance framework using 129 samples (55 eczema and 74 psoriasis). Batch effects were corrected using the ComBat function from the sva package (**Fig. S3A**). Features were initially screened using random forest (RF) and XGBoost models, and subsequently ranked based on SHapley Additive exPlanations (SHAP) values derived from the XGBoost model. The top 10 selected biomarkers for distinguishing eczema and psoriasis were CXCL9, IL-4, IL-17F, IL-17A, CCL8, CSF2, IL-13, VEGFA, CCL11, and CCL19 (**Fig. 2A, Fig. S3B, C**). Of these, CXCL9, IL17F, IL17A, and VEGFA tended to be increased in the psoriasis samples while IL-4, CCL8, IL-13, CCL11, and CCL19 tended to be upregulated in eczema.

**Fig. 2.**
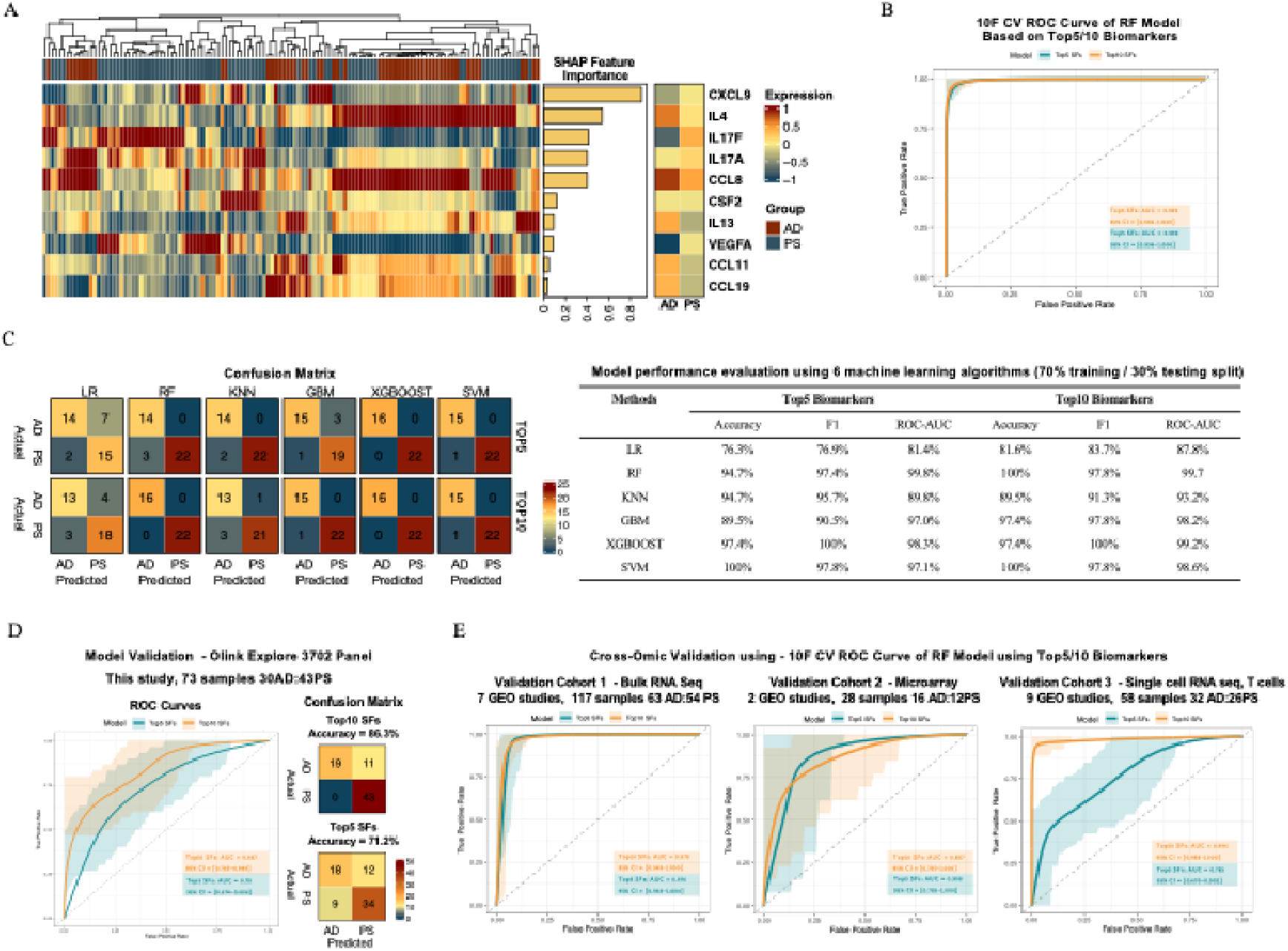
Performance evaluation and validation of machine learning models for psoriasis (PS) and eczema (AD) classification. **(A)** SHAP-based feature importance ranking of the top 10 cytokine biomarkers identified from the Olink panel. **(B)** 10F CV (10-fold cross validation) ROC curve of RF model based on Top5/10 biomarkers. **(C) Left:** Confusion matrices summarizing model predictions from six machine learning algorithms using the top 5 and top 10 selected features (SFs), evaluated using a 70/30 split of the test set (70/30 split). **Right:** Comparison of model performance metrics (accuracy, F1 score, ROC-AUC) across logistic regression (LR), random forest (RF), K-nearest neighbor (KNN), gradient boosting machine (GBM), XGBoost, and support vector machine (SVM) using top 5 and top 10 SFs. **(D)** ROC curves of the best-performing model (RF) using top 5 and 10 SFs, evaluated on an independent validation cohort (Olink Explore 3072 panel). **(E)** Cross-omic validation of the RF model in three external cohorts: bulk RNA-seq, microarray and single-cell RNA-seq T cell data.

To evaluate the predictive performance of the biomarker-based model in this setting, 10-fold cross-validation was performed using a random forest (RF) algorithm. Models built using the top 5 and top 10 biomarkers achieved AUCs of 0.990 and 0.997, respectively (**Fig. 2B**). Subsequently, the data was split into training (70%) and testing (30%) sets to compare six machine learning algorithms, including logistic regression (LR), RF, k-nearest neighbors (KNN), gradient boosting machine (GBM), XGBoost, and support vector machine (SVM). Among these, XGBoost demonstrated superior performance across all evaluation metrics including overall accuracy, F1 score, precision, recall and ROC-AUC and was selected for final model construction (**Fig. 2C, Table S2, Fig. S4A**).

A separate group of DIPS samples from patients with eczema (n=30) and psoriasis (n=43) was analyzed using a much larger panel of protein biomarkers (Olink Explore 3072). While the two Olink platforms utilize a similar protein detection strategy, Explore 3072 uses a next generation sequencing readout with relative quantification, while Target 48 uses a qPCR readout with absolute quantification. To assess the generalizability of the disease classification model trained on the Olink Target 48 dataset, we evaluated its performance using the independent cohort profiled with the Olink Explore 3072. The model achieved robust performance, with area under the curve (AUC) values of 0.78 and 0.867 when using the top 5 and top 10 selected protein biomarkers, respectively. The corresponding classification accuracies were 71.2% and 86.3%, respectively. (**Fig. 2D, Fig. S4B**).

To assess the biological relevance and cross-platform utility of the selected biomarkers independently of the original model, we performed cross-omic validation using publicly available transcriptomic datasets, including bulk RNA sequencing, microarray, and single-cell RNA sequencing data from patients with psoriasis and eczema. First, a bulk RNA-seq data set comprising 54 psoriasis and 63 eczema samples was generated by integrating 7 different published datasets ^22, 23, 24, 25, 26, 27^. A random forest classifier trained solely on the expression of the top five and top ten biomarker genes from the Olink Target 48 cohort achieved AUCs of 0.976 (95% CI: 0.948–1.00) and 0.978 (95% CI: 0.949–1.00), respectively, in 10-fold cross-validation (**Fig. 2E**). Similarly, in a publicly available microarray data set from 12 patients with psoriasis and 16 patients with eczema ^28, 29^, the classifier yielded AUCs of 0.908 (95% CI: 0.785–1.00) and 0.887 (95% CI: 0.765–1.00). Finally, single-cell RNA sequencing data comprising 26,686 T cells was collated and encompassed data from 26 patients with psoriasis and 32 patients with eczema from 9 published studies ^30, 31, 32, 33, 34, 35, 36, 37, 38^. In this data set, the model maintained discriminative power, with AUCs of 0.786 (95% CI: 0.670–0.902) and 0.996 (95% CI: 0.988–1.00) for the top five and top ten biomarkers, respectively (**Fig. 2E**). These findings demonstrate the robustness of the biomarkers identified in the Olink Target 48 cohort across diverse datasets.

### Treatment Response Classifier Development for Eczema

We next examined biomarker heterogeneity within eczema with a particular focus on how it relates to response and nonresponse to commonly used targeted therapies. To do this, we first constructed a treatment response classifier using the Olink Target 48 proteomic data from 31 eczema patients with known therapeutic outcomes: responders to dupilumab (IL-4Rα inhibitor) (IL4_IL13i_R, *n* = 14), non-responders to dupilumab (IL4_IL13i_NR, *n* = 12), and responders to upadacitinib (JAK1 inhibitor) (JAKi_R, *n* = 5). All 5 of the JAKi responders had previously failed dupilumab. Responders were defined as patients achieving an investigator global assessment (IGA) score of 0/1 after 4 months of treatment. Non-responders were defined as patients not achieving this benchmark after 4 months of therapy.

The top 10 markers for distinguishing among these three response patterns included CXCL11, IL-4, CXCL10, IL-17F, IL-27, CSF3, IL-13, IL-7, CCL19 and IL-33 (**Fig. 3A, Fig. S5A**). To assess model robustness, we performed 10-fold cross-validation using an RF classifier trained on the top 10 and top 5 markers, achieving macro AUCs of 0.947 for both, with individual class AUCs above 0.9 (**Fig. 3B**). Considering the limited sample size and class imbalance (only 5 JAKi responders), a stratified 5-fold cross-validation was further conducted, yielding comparable results (**Table S3**). Then, the dataset was split into 70% training and 30% testing subsets to evaluate model generalization. Using the top 10 biomarkers, the RF model achieved a macro-AUC of 0.978, micro-AUC of 0.969, and an overall classification accuracy of 87.5% (**Fig. 3C, Fig. S5B**).

**Fig. 3.**
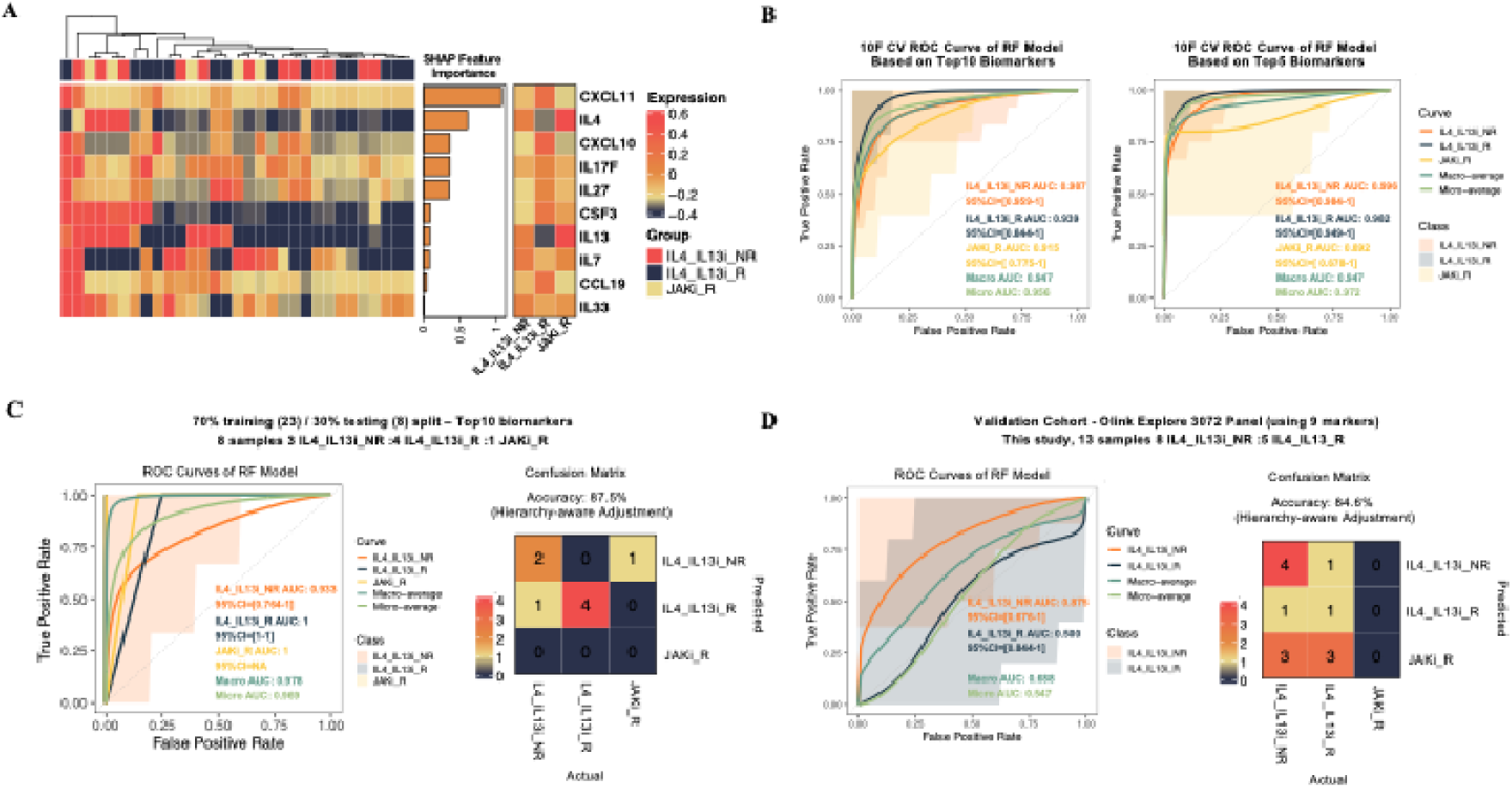
Performance of treatment response classifiers for dupilumab (IL-4/IL-13i) and upadacitinib (JAKi) in eczema. **(A)** SHAP summary plots showing the top 10 most informative cytokine biomarkers correlating with response to dupilumab and upadacitinib and heatmap showing expression of these markers across treatment response groups. R: responder, NR: nonresponder. **(B)** 10-fold cross-validation (10F CV) ROC curves of a random forest (RF) model using the top 10 and top 5 biomarkers. R: responder, NR: nonresponder. **(C)** ROC curves and confusion matrix of the RF model evaluated on a 70:30 train-test split using top 10 biomarkers. R: responder, NR: nonresponder. **(D)** ROC curves and confusion matrix of the RF model using the Olink Explore 3072 data in an independent cohort. R: responder, NR: nonresponder.

To further validate our eczema response classifier, we applied the model to data derived from the Olink Explore 3072 cohort, where therapeutic response data was also available for a subset of patients. IL-27, one of the top 10 eczema response biomarkers, is not part of Olink Explore 3072; thus, a model using the other top 9 biomarkers was used. The model maintained an adjusted classification accuracy of 84.6%, with an AUC of 0.875 for the IL4_IL13i_NR group (**Fig. 3D, Fig. S5C**). The reported accuracy incorporated a hierarchy-aware adjustment, in which predictions labeled as JAKi_R were also considered correct for IL4_IL13i_NR. No dual non-responders were present in the cohort.

Prior studies have demonstrated that elevated *IL4* and *IL13* mRNA levels appear to be associated with favorable response to IL-4/IL-13-targeted therapy (dupilumab); in contrast to *IL4* and *IL13* low/null cases which may rely on other cytokines ^15, 17^. However, we noted in our data, this expected relationship was confounded by a distinct subset of dupilumab non-responders who exhibited IL-4 and IL-13 expression levels substantially exceeding the cohort mean (**Fig. 3A, Fig. S5D**). This interesting observation suggests that similar to patient with negligible expression, patients expressing exceedingly high levels of these biomarkers may not respond well to dupilumab.

We found that the presence of these IL-4/IL-13^very high^ samples did not impair the predictive performance or interpretability of our model, as it is built upon an integrated multi-marker framework rather than reliance on any single marker. SHAP-based feature importance analysis (**Fig. 3A**) confirmed that IL-4 ranked among the top contributors, whereas IL-13 contributed less significantly to model predictions. Importantly, the model incorporates IL-4 expression in the context of other key markers, such as CXCL11 and CXCL10, thereby mitigating the potential bias introduced by a few IL-4/IL-13^very high^ samples.

The robustness of our model was further supported by its consistent predictive accuracy in an independent validation cohort (**Fig. 3D**), underscoring its generalizability despite this apparent biologic complexity and a potential U-shaped relationship with IL-4 and IL-13 levels in terms of dupilumab response.

### Treatment Response Classifier Development for Psoriasis

Next, we focused on biomarker heterogeneity in the Target 48 psoriasis cohort with the goal of exploring biomarkers correlating with treatment response. Although patients were treated with a number of different therapies, we first focused on samples from 14 patients treated with apremilast (PDE4 inhibitor). Identifying apremilast responders could be particularly valuable because in contrast to the other psoriasis medications we evaluated, apremilast is an oral medication that is not thought to carry some of the risks that have been associated with other injectable biologics. Of the apremilast sample cohort, there were 6 responders and 8 nonresponders (**Fig. 4A, Fig. S6C**). Response/nonresponse was defined in a manner analogous to the eczema patients. Analyses were also performed for IL-17A inhibition (secukinumab and ixekizumab) (IL17i, **Fig. S6A**), IL-23p19 inhibition (guselkumab and risankizumab) responsiveness (IL23i, **Fig. 4B, Fig. S6B**), and IL-12/23p40 inhibition (ustekinumab) (IL12_23i, **Fig. S6D**); however, the number of patients in each of these groups was relatively low, so we considered these analyses as preliminary.

**Fig. 4.**
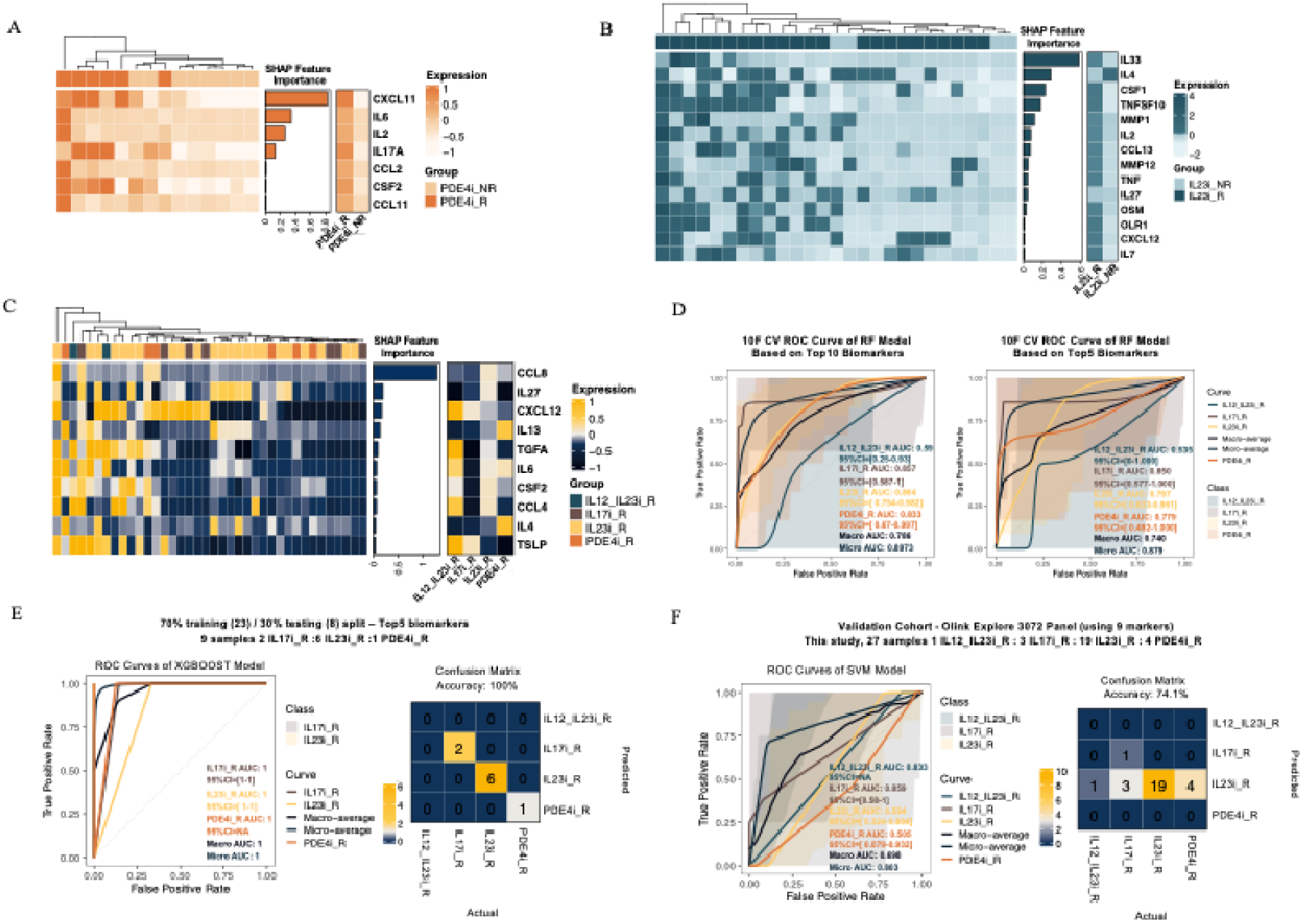
Performance of treatment response classifiers to ustekinumab (IL-12/IL-23), IL-17A inhibition, IL-23 inhibition), and apremilast (PDE4) in psoriasis. **(A)** SHAP summary plots showing the selected important cytokine biomarkers for distinguishing responders to apremilast and heat map showing levels of expression of the markers across response subgroups. R: responder, NR: nonresponder. **(B)** SHAP summary plots showing the selected important cytokine biomarkers for distinguishing responders to IL-23 inhibition and heat map showing levels of expression of the markers across response subgroups. R: responder, NR: nonresponder. **(C)** SHAP summary plots showing the top 10 most informative cytokine biomarkers correlating with response patterns to all drugs and heat map showing levels of expression of the markers across response subgroups. R: responder, NR: nonresponder. **(D)** ROC curves from 10-fold cross-validation of a random forest model based on the top 10 biomarkers. R: responder, NR: nonresponder, 10F CV: 10-fold cross validation. **(E)** ROC curves and confusion matrix to evaluate the XGBoost model using a 70:30 training/testing split. R: responder, NR: nonresponder. **(F)** ROC curves and confusion matrix in the validation Olink Explore 3072 panel cohort. R: responder, NR: nonresponder.

Considering potential clinical utility, we first constructed a classifier to evaluate response to all of these different psoriasis medications using the Olink Target 48 panel. This group consisted of 31 samples from psoriasis patients with annotated treatment outcomes. The top 10 markers selected via SHAP-based interpretation were CCL8, IL-27, CXCL12, IL-13, TGF-α, IL-6, CSF2, CCL4, IL-4, and TSLP (**Fig. 4C**). Model performance was first assessed using 10-fold cross-validation, yielding a macro-AUC of 0.786 and micro-AUC of 0.897 across the four drug classes (PDE4, IL-17A, IL-23p19, and IL-12/23p40) (**Fig. 4D**). A subsequent 70:30 train-test split confirmed generalizability, with an accuracy of 100% in the test set and per-class AUCs ranging from 0.779 to 0.850 **(Fig. 4E, Fig. S7A, B**). To evaluate external validity, the classifier was applied to an independent validation cohort of psoriasis samples with annotated treatment outcomes (n = 27) profiled using the Olink Explore 3072 panel. A reduced model based on nine overlapping markers achieved 74.1% accuracy and a macro-AUC of 0.698 (**Fig. 4F, Fig. S7C, D**). In the future, we expect improved model performance as data from additional samples are integrated into the model.

### Development of the DIPS-Derm Web Platform for Clinical Decision Support

In order to facilitate implementation of the models in a research setting outside of our institution, external validation, ongoing data collection, and expansion of the approach, we developed an interactive web-based platform, DIPS-Derm (https://dips-derm.onrender.com). DIPS-Derm is based on the best-performing models and datasets identified in this study. The platform allows users to upload proteomic (Olink) profiles or gene expression profiles (bulk RNA and single cell RNA sequencing) to obtain automated predictions for disease classification (eczema vs. psoriasis) and individualized treatment response (e.g. dupilumab R and NR and apremilast R and NR). The platform is presently available for research purposes (**Fig. 5**) and allows users to include key nonidentifying demographic information and clinical features of disease.

**Fig. 5.**
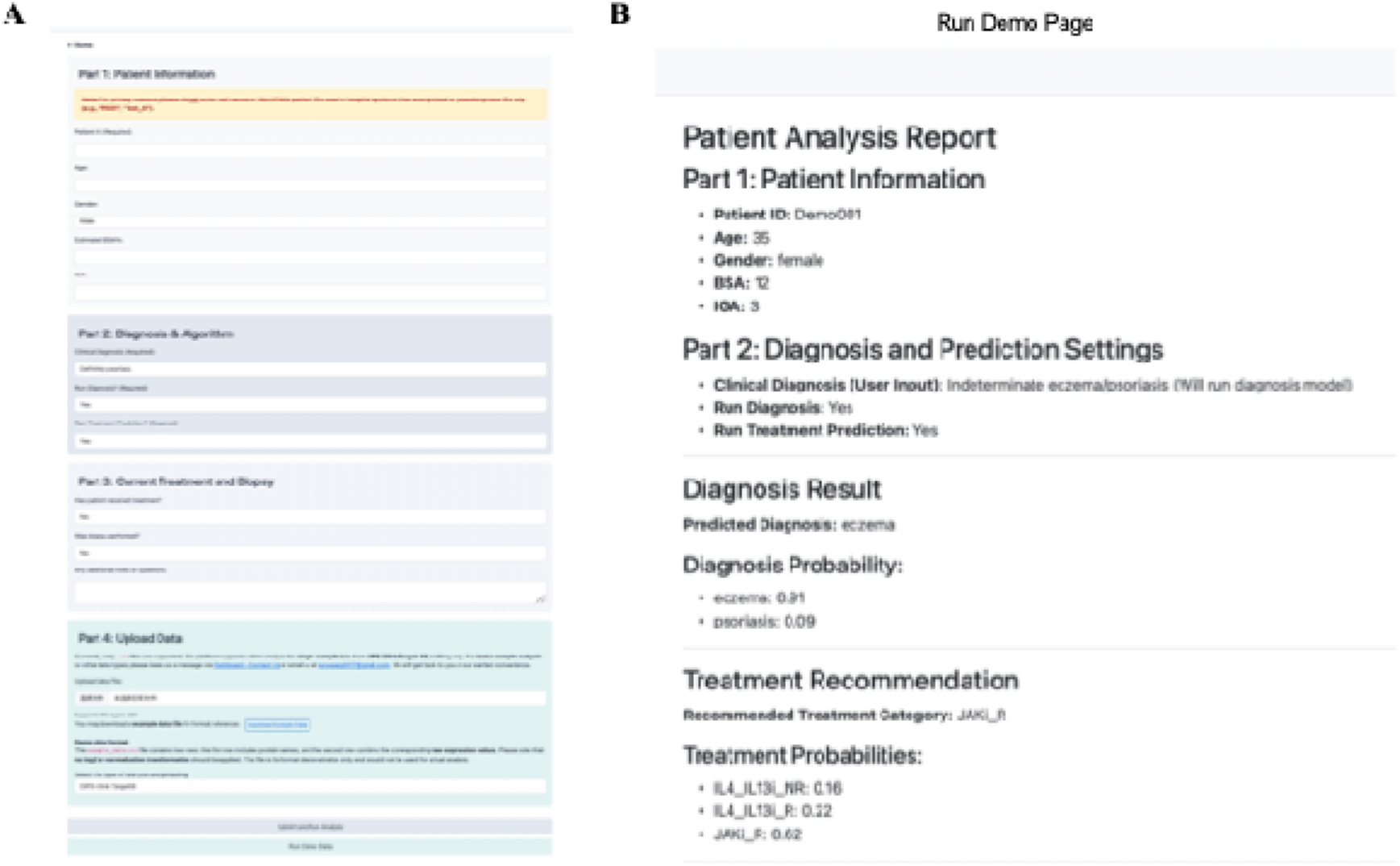
User interface and demonstration of the DIPS-Derm web platform. **(A)** Screenshot of the DIPS-Derm platform interface after user login. **(B)** Demonstration of the output using example data. R: response, NR: nonresponse.

## Discussion

Personalized medicine for inflammatory skin disease has been difficult to achieve but holds tremendous promise. Even when a skin biopsy is performed, some cases of eczema and psoriasis may still be difficult to distinguish ^29, 39, 40^. Outside of very few settings, probing the unique immune polarization of any individual patient, as part of a typical clinical workflow, is not possible. These factors make treatment selection for these patients inefficient and trial-and-error based, resulting in frequent treatment switches. Even in cases in which the diagnosis of eczema or psoriasis is clear, treatment selection remains almost entirely empiric (trial-and-error) without the ability to incorporate personalized immunologic information into therapeutic decision making.

Here, we present DIPS and related machine learning approaches as a potential application of personalized medicine in dermatology. We found the models built using data derived from the DIPS specimens to perform very well in various tasks including differentiating between psoriasis and eczema and in predicting therapeutic response and nonresponse in some settings. We expect that adding data from additional samples over time will continue to improve performance of the models. The models have been built into a web-based platform so that data may be uploaded by other researchers to facilitate collection of additional data. We are also beginning to evaluate the feasibility of incorporating additional clinical data in the model to increase value and utility.

Other personalized medicine approaches for inflammatory skin diseases have been described. For example, Mind.Px^TM^ from Mindera Health has been developed for treatment recommendations for patients with psoriasis. Mind.Px^TM^ utilizes a dermal biomarker patch, which extracts mRNA for subsequent gene expression profiling ^41^. Castle Biosciences has also reported a program in treatment selection for atopic dermatitis based on gene expression profiling, but to our knowledge, no data has been published. Other efforts in this space have included RNA-seq on small punch biopsies ^42^ and sc-RNA-seq from dissociated skin biopsies ^43^. In our own work, we have used cytokine staining in tissue biopsies for similar purposes ^42^; however, a biopsy is necessary, and the technique is inherently low-plex in terms of the number of biomarkers that can be reasonably evaluated.

The first potential advantage to DIPS is the ease of sample collection, which is accomplished in under 1 minute and is faster than standard (e.g. skin biopsy) or other approaches ^44^. This suggests that clinicians would have a low bar to integrate sample collection into their clinical workflow. Moreover, a recent survey study suggested that dermatologists would be willing to use a personalized medicine test, would alter first-line treatment based on such a test, and would use test results in the insurance approval process ^45^. Further, patients have found this minimally invasive sample collection approach to be exceedingly well tolerated. Once the samples have been collected there is minimal downstream workflow; the samples are ready for analysis upon collection.

Another potential advantage is collection of material from full-thickness epidermis, where the majority of relevant biomarkers appear to be present in eczema and psoriasis ^13^ coupled with detection of biomarkers at the protein, as opposed to the mRNA, level. Last, analysis with our approach is fully quantitative, suggesting that in the future, reference values for key biomarkers could be generated.

Limitations of the work include the retrospective and single-center nature of the analysis and relatively low sample numbers, particularly for treatment response evaluation in psoriasis. Nonetheless, we believe that this work helps set the foundation for a platform approach with the potential to realize personalized medicine for diagnosis and treatment of inflammatory skin disease.

## Methods

### Sample collection

We utilized a previously described surfactant combination as the basis for the DIPS procedure ^20^. This surfactant consists of 0.25% Laureth-4 (Brij-30 and polyoxyethylene 4 lauryl ether); 0.25% DPS-30 (N-decyl-N,N-dimethyl-3-ammonio-1-propanesulfonate); 15% boric acid/potassium chloride buffer, pH 9.5; 0.25% polysorbate-20, NF grade; 0.5% hydroxypropylcellulose, NF; 0.5% diazolidinyl urea; 0.25% phenylethyl alcohol] now termed Clearista Retexturizing Gel (Skincential Sciences, San Francisco). Skin was swabbed using a microfiber cosmetic applicator and approximately 40µL of surfactant. Swabbing was performed by rubbing the surfactant-coated applicator tip in a back-and-forth motion for 45 seconds, applying moderate pressure, over a 2mm-by-1mm rectangular area on skin. For psoriasis and eczema sample collection, lesional skin was utilized. The tip of the swab was then inserted into an Eppendorf tube containing 130µL of phosphate buffered saline and a protease inhibitor cocktail (Sigma-Aldrich #11836153001), let sit for 1 minute, and then rubbed against the interior of the tube to dislodge any additional attached material. Samples were then immediately placed on wet ice for transport and later stored at −80°C prior to analysis.

### Protein biomarker measurement

The discovery cohort samples were analyzed using Olink Target 48 Cytokine Panel (Olink Proteomics, Uppsala, Sweden), which provides absolute quantification of 45 cytokine and chemokine proteins using a proximity extension assay (PEA)-based technique coupled with real-time qPCR. A validation cohort was analyzed using the Olink Explore 3072 Panel which provides relative quantification of 3072 proteins. This assay also utilizes a PEA-based technique but is instead coupled with NGS readouts. The manufacturer’s instructions were followed. Deidentified samples were run at Vanderbilt University in the High-Throughput Biomarker Core.

### Patient selection

Patients were enrolled from a single outpatient dermatology practice at a tertiary academic referral center. Inclusion criteria required patients to be aged ≥18 years with a clinical and/or histopathologic diagnosis of psoriasis or eczema confirmed by an experienced board-certified dermatologist. The cohort included cases with both classical and non-classical morphologies. Clinical severity was assessed using BSA and IGA (psoriasis) or validated IGA for atopic dermatitis (vIGA-AD). For each participant, at least one lesional skin sample was collected, along with basic demographic information, treatment history, and standardized clinical photographs of the sampled lesion.

### Machine learning model construction and validation for Olink data

Machine learning analyses were conducted using both R (version 4.2.0) and Python (version 3.8). Proteomic feature selection was first performed using the random forest (RF) models were implemented in R using the randomForest (v4.7-1) package. Features were ranked by Gini importance, and the top 20 proteins were selected for subsequent modeling. Extreme gradient boosting (XGBoost) models were constructed using the xgboost (v1.6.0.1) package. Feature importance was assessed based on SHapley Additive exPlanations (SHAP) values derived from XGBoost models, using the SHAPforxgboost (v0.1.3) package. Top-ranked biomarkers were selected for model construction.

Model training, testing, and performance evaluation were performed in R using the xgboost (v1.6.0.1) package. A 7:3 train-test split was applied to the discovery cohort (Olink Target 48 Cytokine cohort). Six machine learning algorithms were evaluated, including logistic regression (LR), RF, k-nearest neighbors (KNN), gradient boosting machine (GBM), extreme gradient boosting (XGBoost), and support vector machine (SVM). Hyperparameters were optimized via grid search where applicable.

Model performance was assessed based on overall accuracy, F1 score, precision, recall, and area under the receiver operating characteristic curve (ROC-AUC). 10-fold cross-validation (10FCV) was additionally performed to internally validate model robustness prior to external validation. Validation for the eczema versus psoriasis classification model was performed across three independent cohorts without retraining: the Olink Explore 3072 cohort, combined GEO microarray datasets (GSE185764 and GSE182740), and a single-cell RNA-sequencing T cell dataset (see below).

### External validation data collection

#### Combined bulk RNA sequencing cohort

Publicly available bulk RNA-seq datasets were retrieved from the Gene Expression Omnibus (GEO) database for cross-omic validation. 7 independent datasets were selected: GSE176279, GSE65832, GSE121212, GSE249936, GSE67785, GSE41745, and GSE232127^22, 23, 24, 25, 26, 27^. Raw gene count matrices were downloaded from the GEO portal. For each dataset, expression values corresponding to the selected biomarker genes (*CXCL9*, *IL4*, *IL17F*, *IL17A*, *CCL8*, *CSF2*, *IL13*, *VEGFA*, *CCL11*, *CCL19*) were extracted and reformatted into a sample-by-gene matrix. Samples were manually labeled as either eczema or psoriasis (PS) based on the provided metadata. To enable joint analysis, expression matrices from all datasets were merged by gene symbol, and sample-level log2 transformation was applied. Batch effects across the seven datasets were corrected using the ComBat function from the sva R package (version 4.2.0), with disease group specified as a covariate to preserve biological variation. The resulting harmonized dataset was used for downstream classification analysis.

#### Combined GEO microarray cohort

Publicly available microarray datasets were retrieved from the Gene Expression Omnibus (GEO) database for external validation. Two independent datasets were selected: GSE182740 (Affymetrix Human Genome U133 Plus 2.0 Array, GPL570 platform) and GSE185764 (Affymetrix Human Transcriptome Array 2.0, GPL17586 platform). For GSE182740, raw microarray files (CEL.gz) were processed using the affy (v1.74) R package^28, 29^. Robust Multi-array Average (RMA) normalization, including background correction, quantile normalization, and probe summarization, was applied. Probe sets were mapped to gene symbols using the hgu133plus2.db annotation (v3.31.0) package. Probes without gene symbol annotations were excluded, and for genes represented by multiple probes, mean expression values were computed. For GSE185764, raw CEL files were processed using the oligo (v1.60.0) R package with RMA normalization. Gene annotation was performed using the hta20transcriptcluster.db (v8.8.0) package, and similar to GSE182740, probes lacking gene symbols were removed and multiple probes corresponding to the same gene were aggregated by averaging. Following preprocessing, expression matrices from both datasets were harmonized by selecting the intersecting set of genes present in the discovery cohort. Batch effects across datasets were corrected using the ComBat function from the sva R package, with disease group included as a covariate to preserve biological variability.

#### Single cell cohort

Publicly available single-cell RNA sequencing (scRNA-seq) datasets were retrieved from the Gene Expression Omnibus (GEO) database, including GSE183047, GSE151177, GSE158432, GSE153760, GSE213849, GSE228421, GSE230842, GSE180885, and GSE230575^30, 31, 32, 33, 34, 35, 36, 37, 38^. Raw or processed gene expression matrices were imported into R and analyzed using the Seurat package (v5.3.0). Standard quality control procedures were applied according to each dataset’s original processing criteria. T cells were identified based on expression of canonical T cell markers (*CD3E*, *CD3D*, and *CD3G*), and cells expressing at least one of these markers above a defined threshold were retained for further analysis. After extraction, T cells from different datasets were merged into a single Seurat object. Batch effects among datasets were corrected using the Harmony package (v1.2.3) to align cells across different studies while preserving biological variation. All the processed GEO datasets were reserved exclusively for external validation and were not used during model training, feature selection, or hyperparameter tuning.

#### Study Approval

This study was reviewed and approved by the Yale University institutional review board (#2000033233). Written informed consent was received from all patients prior to inclusion in the study.

## Data Availability

All data produced in the present study are available upon reasonable request to the authors

## Acknowledgments

This publication was made possible by the Yale School of Medicine Fellowship for Medical Student Research and The Richard K. Gershon, M.D., Endowed Medical Student Research Fellowship (MJM). AMW and SLS were supported by the National Institute of Diabetes and Digestive and Kidney Diseases of the National Institutes of Health under Award Number T35DK104689. WD was supported by the LEO Foundation. The content is solely the responsibility of the authors and does not necessarily represent the official views of the National Institutes of Health.

## Author contributions

JMC and WD conceptualized and designed the research studies and enrolled patients in the study. MJM optimized the DIPS protocol. YW designed and visualized the AI framework, developed and tested machine learning models, performed diagnostic classifier construction, and analyzed multi-modal data. TC assisted with machine learning model testing. GC and PE conducted experiments. MJM, GC, PE, AMW, and SLS acquired clinical and experimental data. WD provided funding. KS provided the Clearista reagent. YW, MJM, GC, PE, AMW, SLS, JMC, and WD contributed to manuscript writing and critical.

## Data and materials availability

The raw Olink data and supporting code are available from the authors upon reasonable request.

