## Supplementary Figures for "Non-invasive epidermal proteomics and machine learning permits molecular subclassification of psoriasis and eczematous dermatitis"

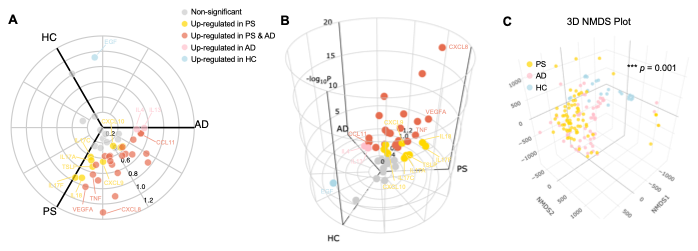

**Fig. S1. DIPS enables quantitative epidermal protein biomarker profiling and discrimination of psoriasis (PS), eczema (AD), and healthy control (HC) skin.**

**(A)** Two-dimensional polar plot depicting the relative levels of biomarkers using data from the Olink Target 48 Cytokine panel. Each point represents a biomarker, positioned according to relative fold change between groups. Significantly upregulated markers are color-coded accordingly, with nonsignificant proteins shown in gray.

**(B)** Three-dimensional representation of the same dataset illustrating the magnitude and significance (–log₁₀ P) of protein expression differences among the groups.

**(C)** Three-dimensional nonmetric multidimensional scaling (NMDS) plot based on protein levels showing distinct clustering of each group (PERMANOVA, p = 0.001).

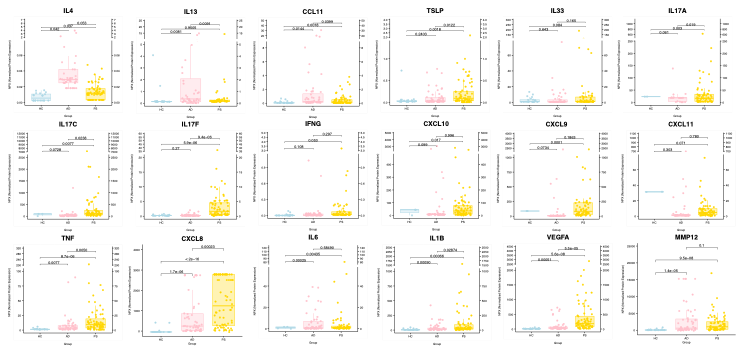

**Fig. S2. Protein biomarker levels in psoriasis (PS), Eczema (AD), and healthy controls (HC).** Boxplots displaying absolute concentrations (pg/μL) of representative cytokine, chemokine, and other markers using data from the Olink Target 48 Cytokine panel. Each dot represents an individual sample, and boxes indicate interquartile ranges and median (horizontal line).

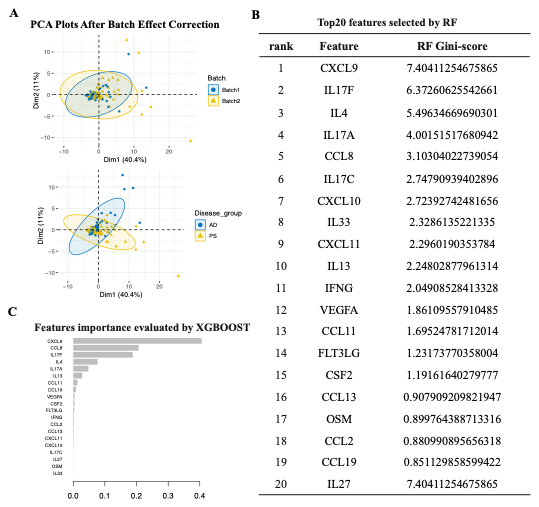

**Fig. S3. Identification of key protein biomarkers distinguishing psoriasis (PS) and eczema (AD) using machine learning–based feature selection.**

**(A)** Principal component analysis (PCA) plots of Olink Target 48 cytokine data after batch effect correction.

**(B)** Top 20 protein features ranked by Random Forest (RF) Gini importance score.

**(C)** Feature importance ranking derived from the XGBoost model.

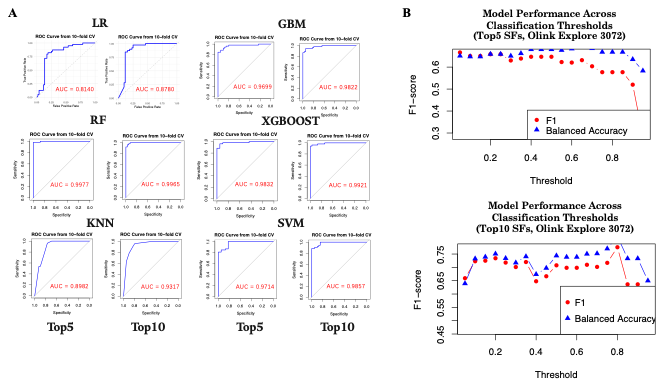

**Fig. S4. Performance assessment of model to distinguish psoriasis and eczema.**

**(A)** Receiver operating characteristic (ROC) curve from 10-fold cross-validation (10F CV) demonstrating the classification performance of six machine learning algorithms—logistic regression (LR), random forest (RF), k-nearest neighbors (KNN), gradient boosting machine (GBM), XGBoost, and support vector machine (SVM). Models were trained using the top five or top ten selected biomarkers from the Olink Target 48 Cytokine dataset.

**(B)** External validation of the psoriasis versus eczema classifier using an independent DIPS cohort profiled with Olink Explore 3072. Model performance was evaluated across varying classification thresholds using top five (upper panel) and top ten (lower panel) biomarkers. F1 score (red) and balanced accuracy (blue) are plotted, showing optimal thresholds corresponding to AUCs of 0.78 and 0.867, respectively. SF: selected feature.

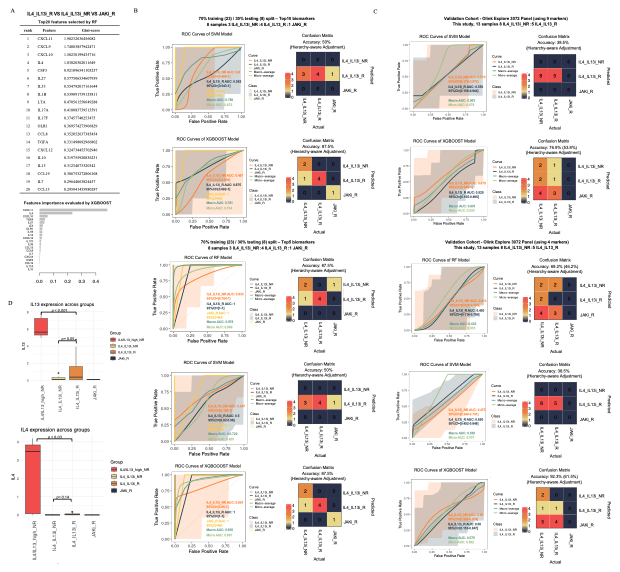

**Fig. S5. Development and validation of a classifier for treatment response in eczema.**

**(A)** Feature selection for treatment response classification using Olink Target 48 Cytokine data from eczema patients treated with dupilumab (IL-4/IL-13) or upadacitinib (JAK). The top 20 protein markers were ranked by Random Forest (RF) Gini importance (upper panel) and XGBoost feature importance (lower panel). R: responder, NR: nonresponder.

**(B)** Model performance on internal validation using a 70% training and 30% testing split. ROC curves and confusion matrices are shown for RF, SVM, and XGBoost classifiers trained on the top 10 or top 5 features. R: responder, NR: nonresponder.

**(C)** External validation using the Olink Explore 3072 dataset. Because IL27 is not included in the Explore panel, a reduced nine-marker (top 9) and four-marker (top 4) model were evaluated to assess cross-platform generalizability. Model performance is eveluated using ROC curves and confusion matrices for RF, SVM, and XGBoost classifiers. Reported accuracy reflects a hierarchy-aware adjustment, in which predictions labeled as JAKi_R were also considered correct for IL4_IL13i_NR. Values in parentheses indicate the absolute accuracy. R: responder, NR: nonresponder.

**(D)** Boxplots showing IL-13 (top) and IL-4 (bottom) protein expression across response groups. R: responder, NR: nonresponder.

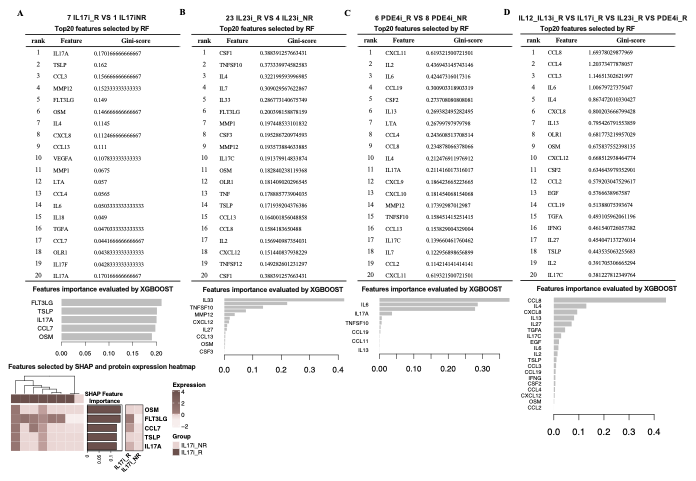

**Fig. S6. Identification of biomarkers associated with treatment response in psoriasis.** Feature selection and model interpretation for psoriasis treatment response classifiers were trained using Olink Target 48 Cytokine data. Each panel shows the top 20 features ranked by Random Forest (RF) Gini importance (upper tables) and corresponding feature importance evaluated by XGBoost (middle bar plots).

**(A)** IL-17A inhibitor (IL17i) responder (R, n = 7) versus non-responder (NR, n = 11) comparison.

**(B)** IL-23 inhibitor (IL23i) responder (R, n = 23) versus non-responder (NR, n = 14) comparison.

**(C)** Apremilast (PDE4) responder (R, n = 6) versus non-responder (NR, n = 8) comparison.

**(D)** Cross-pathway comparison of ustekinumab (IL12/IL23i)_R, IL17i_R, IL23i_R, and apremilast (PDE4i_R) groups identified shared and distinct proteomic signatures associated with response to different drug classes. R: responder, NR: nonresponder.

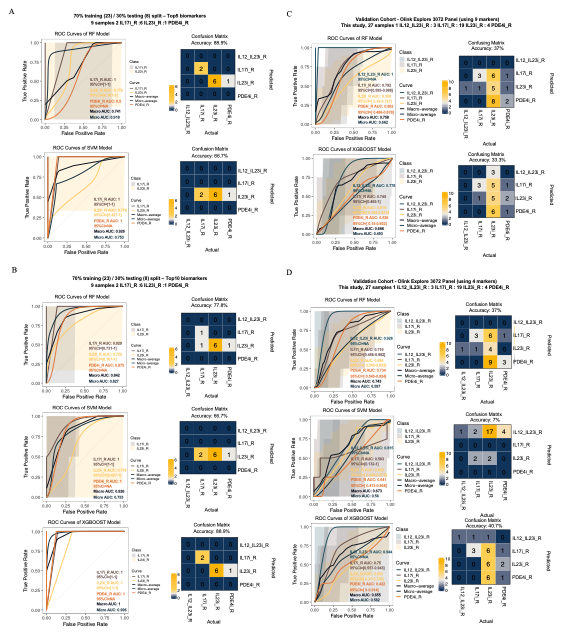

**Fig. S7. Construction and validation of a multidrug response classifier in psoriasis.**

(**A-B**) Model performance on internal validation using a 70% training and 30% testing split. ROC curves and confusion matrices are shown for RF, SVM, and XGBoost classifiers trained on the **(A)** top 5 or **(B)** top 10 features. R: responder, NR: nonresponder.

**(C–D)** External validation using an independent psoriasis cohort (n = 27) profiled with the Olink Explore 3072 panel. Due to differences in protein coverage between the two Olink platforms, a reduced model using **(C)** nine overlapping markers (excluding IL27) and a minimal **(D)** four-marker version were evaluated to assess generalizability. ROC curves and confusion matrices are shown for RF, SVM, and XGBoost models. R: responder, NR: nonresponder.
