## Supplementary Tables for "Non-invasive epidermal proteomics and machine learning permits molecular subclassification of psoriasis and eczematous dermatitis"

**Table S1** Tolerability Evaluation of DIPS Procedure using Local Tolerability Score^1^

| Patient ID | pre collection | | | during collection | | | 10 mins after collection | | |
| --- | --- | --- | --- | --- | --- | --- | --- | --- | --- |
|  | burning/  stinging | pruritus | erythema | burning/  stinging | pruritus | erythema | burning/  stinging | pruritus | erythema |
| P1 | 0 | 0 | 0 | 1 | 0 | 0 | 0 | 0 | 1 |
| P2 | 0 | 0 | 0 | 1 | 0 | 0 | 1 | 0 | 1 |
| P3 | 0 | 0 | 0 | 0 | 0 | 2 | 0 | 0 | 2 |
| P4 | 0 | 0 | 0 | 1 | 0 | 1 | 0 | 0 | 0 |
| P5 | 0 | 0 | 0 | 1 | 0 | 1 | 0 | 0 | 1 |
| P6 | 0 | 0 | 0 | 0 | 0 | 1 | 0 | 0 | 0 |
| **Mean** | **0** | **0** | **0** | **0.7** | **0** | **0.8** | **0.2** | **0** | **0.8** |

^1^L.T. Zane, M.H. Hughes, S. Shakib, Tolerability of Crisaborole Ointment for Application on Sensitive Skin Areas: A Randomized Double-Blind, Vehicle-Controlled Study in Healthy Volunteers. *Am J Clin Dermatol* **17**, 519-526 (2016).

Possible scores for each domain: 0 (none), 1 (mild), 2 (moderate), or 3 (severe)

**Table S2** Diagnostic classifier performance for distinguishing eczema and psoriasis using Olink top 5 and top 10 biomarkers (7:3 training:test ROC).

| Method | Top5 | | | | | Top10 | | | | |
| --- | --- | --- | --- | --- | --- | --- | --- | --- | --- | --- |
|  | OA | Pre | Rec | F1 | AUC | OA | Pre | Rec | F1 | AUC |
| LR | 76.3% | 68.2% | 88.2% | 76.9% | 81.4% | 81.6% | 81.8% | 85.7% | 83.7% | 87.8% |
| RF | 94.7% | 100% | 95% | 97.4% | 99.8% | 100% | 100% | 95.7% | 97.8% | 99.7% |
| KNN | 94.7% | 100% | 91.7% | 95.7% | 89.8% | 89.5% | 95.5% | 87.5% | 91.3% | 93.2% |
| GBM | 89.5% | 86.4% | 95% | 90.5% | 97% | 97.4% | 100% | 95.7% | 97.8% | 98.2% |
| XGBOOST | 97.4% | 100% | 100% | 100% | 98.3% | 97.4% | 100% | 100% | 100% | 99.2% |
| SVM | 100% | 100% | 95.7% | 97.8% | 97.1% | 100% | 100% | 95.7% | 97.8% | 98.6% |

OA: overall accuracy

Pre: precision

Rec: Recall,

AUC: Area Under the Curve

**Table S3** Performance of the eczema treatment response classifier under stratified 5-fold cross-validation

| Features | Macro AUC | Micro AUC | Class | Precision | Recall | Specificity | Balanced Accuracy | F1 |
| --- | --- | --- | --- | --- | --- | --- | --- | --- |
| Top10 | 0.929 | 0.921 | IL4_IL13i_R | 0.82 | 1.00 | 0.82 | 0.91 | 0.90 |
|  |  |  | IL4_IL13i_NR | 0.91 | 0.83 | 0.95 | 0.89 | 0.87 |
|  |  |  | JAKi_R | 1.00 | 0.60 | 1.00 | 0.80 | 0.75 |
| Top5 | 0.947 | 0.944 | IL4_IL13i_R | 0.87 | 0.93 | 0.88 | 0.91 | 0.90 |
|  |  |  | IL4_IL13i_NR | 0.85 | 0.92 | 0.89 | 0.91 | 0.88 |
|  |  |  | JAKi_R | 1.00 | 0.60 | 1.00 | 0.80 | 0.75 |

Per-class performance metrics (Precision, Recall, Specificity, Balanced Accuracy, and F1) and overall macro/micro AUCs (out-of-fold, OOF) are reported for models trained on the top 10 and top 5 biomarker sets.
